# Estimating Ejection Fraction from the 12 Lead ECG among Patients with Acute Heart Failure

**DOI:** 10.1101/2024.03.25.24304875

**Authors:** Sunita Pokhrel Bhattarai, Dillon J Dzikowicz, Ying Xue, Robert Block, Rebecca G. Tucker, Shilpa Bhandari, Victoria E Boulware, Breanne Stone, Mary G Carey

## Abstract

**Background:** Identifying patients with low left ventricular ejection fraction (LVEF) in the emergency department using an electrocardiogram (ECG) may optimize acute heart failure (AHF) management. We aimed to assess the efficacy of 527 automated 12-lead ECG features for estimating LVEF among patients with AHF.

**Method:** Medical records of patients >18 years old and AHF-related ICD codes, demographics, LVEF %, comorbidities, and medication were analyzed. Least Absolute Shrinkage and Selection Operator (LASSO) identified important ECG features and evaluated performance.

**Results:** Among 851 patients, the mean age was 74 years (IQR:11), male 56% (n=478), and the median body mass index was 29 kg/m^2^ (IQR:1.8). A total of 914 echocardiograms and ECGs were matched; the time between ECG-Echocardiogram was 9 hours (IQR of 9 hours); ≤30% LVEF (16.45%, n=140). Lasso demonstrated 42 ECG features important for estimating LVEF ≤30%. The predictive model of LVEF ≤30% demonstrated an area under the curve (AUC) of 0.86, a 95% confidence interval (CI) of 0.83 to 0.89, a specificity of 54% (50% to 57%), and a sensitivity of 91 (95% CI: 88% to 96%), accuracy 60% (95% CI:60 % to 63%) and, negative predictive value of 95%.

**Conclusions:** An explainable machine learning model with physiologically feasible predictors may be useful in screening patients with low LVEF in AHF.

**Clinical Perspective:** 

**What is new?:**

- Among 527 ECG features, 42 were important in estimating ≤30% reduced left ventricular ejection fraction (LVEF), showing the model’s high diagnostic accuracy (AUC of 0.86).
- The model exhibits exceptional sensitivity (91%) in predicting ≤30% LVEF
- ECG-derived metrics offer the potential for early detection of reduced LVEF, especially in settings with limited advanced diagnostic tools.

**What are the clinical implications?:**

- Enhanced diagnostic accuracy allows for the earlier detection of reduced LVEF through ECG analysis, which is critical in an environment where an echocardiogram is unavailable.
- ECG features enable patient risk stratification for reduced LVEF, facilitating targeted management and optimization of healthcare resources.
- The findings underscore the importance of integrating ECG features into AI-based diagnostic models for rapid, accurate LVEF estimation, supporting more informed clinical decisions and enabling effective remote patient monitoring.

## Introduction

Heart failure (HF) is a complex syndrome arising from either functional or structural impairment of ventricles, characterized by symptomatic left ventricular dysfunction. The impairment in left ventricle function causes reduced efficacy in pumping blood, thereby instigating a cascade of adverse consequences, including pulmonary edema, arrhythmias, and increased risk of sudden cardiac death, especially in low ejection fractions ^1^. The United States recorded 139.8 million emergency department (ED) visits in 2023 ^2^, and 4% were patients with acute heart failure (AHF) ^3^. Acute heart failure is defined by a rapid or gradual onset of symptoms and signs of HF, leading patients to seek urgent medical attention with unplanned hospitalization or an ED visit ^4,5^. AHF is an unstable condition, and the severity of the symptoms can increase; thus, identifying low left ventricular ejection fraction (LVEF) quickly at ED could prevent deterioration of myocardial performance and prevent early death ^6^. Despite the critical significance of LVEF measurement, clinicians frequently report delays in obtaining these measurements primarily because specialized imaging modalities and high-skill intensive tests such as echocardiography or cardiac MRI are required ^4,5,7,8^. Specialized procedures demand highly skilled resources and are not universally accessible across all healthcare settings.^8^ Addressing this challenge underscores the need for a readily accessible diagnostic tool to estimate LVEF where immediate specialized imaging is not available. Easily accessible diagnostic tools might significantly mitigate delays and accelerate the initial triage of patients with HF.

The 12-lead electrocardiogram (ECG) is a universal first-line diagnostic tool to assess cardiac activity, such as structural changes and conduction disorders. It is widely accessible, provides instant results, and is inexpensive ^9,10^. Different studies demonstrated that ECG features correlate with different categories of LVEF, including temporal ECG intervals, ECG vectors, and other ECG morphologies in research and clinical practice ^10,11^. In recent years, ECG has been widely used to predict LVEF using artificial intelligence ^8,12–14^. Kwon et al.^13^ applied deep learning using ECG; they performed echocardiography within four weeks to predict HF with reduced EF (≤40%) and with mid-range to reduced EF(≤50%), achieving an area under the curve (AUC) of 0.843 in internal validation and 0.889 in external validation and outperforming other models of logistic regression and random forest algorithm. A study from the Mayo Clinic with ECG to EF window of two weeks trained deep learning models to identify patients with EF≤35% with a high AUC of 0.932 using ECG alone ^8^. Observing previous studies findings, it has been suggested that deep learning models (DLM) could estimate low LVEF using ECG features.

Despite the impressive accuracy achieved using deep learning in several studies, it is important to interrogate these algorithms to understand how they work, including their transparency, reproducibility, and dissemination in clinical practices^15^. It remains challenging to infer the electrophysiological mechanisms underlying this association precisely. Therefore, it remains a significant missed opportunity in clinical practice. An explainable model is needed to estimate reduced LVEF. Accordingly, we aimed to assess the efficacy of 527 automated, artificial intelligence-based 12-lead ECG features for estimating reduced LVEF (≤30%) among patients with AHF using an explainable LASSO machine-learning algorithm to identify key ECG features and evaluate model performance.

## Methods

### Study Design, Data Source, and Population

The University of Rochester’s institutional review board approved this study. We retrospectively collected data from electronic health records (Epic Systems, Verona, Wisconsin) among patients with AHF treated in the EDs and hospitalized between January 1, 2016, and December 31, 2019. This time frame was established based on two key considerations: first, the official adoption of the ICD-10 code in the hospital occurred in 2016, and second, to mitigate the historical data threats associated with COVID-19.

### Inclusion and Exclusion Criteria

We selected patients above 18 years old who had encountered ED visits with a principal diagnosis of HF ICD-10 codes. These patients also had recorded LVEF measurements and ECGs within 24-hour window. To refine our selection, we excluded HF patients who had a previous diagnosis of myocarditis, pericarditis, and endocarditis within 30 days of previous HF hospitalization, and HF patients who presented with acute coronary syndromes because of potential ECG changes unrelated to AHF ^16^. We also excluded patients using left ventricular assist devices (n=20), pacemakers (n=74) and missing an ECG (n=80) yielding a total of 851 unique patients (Figure 1).

**Figure 1.**
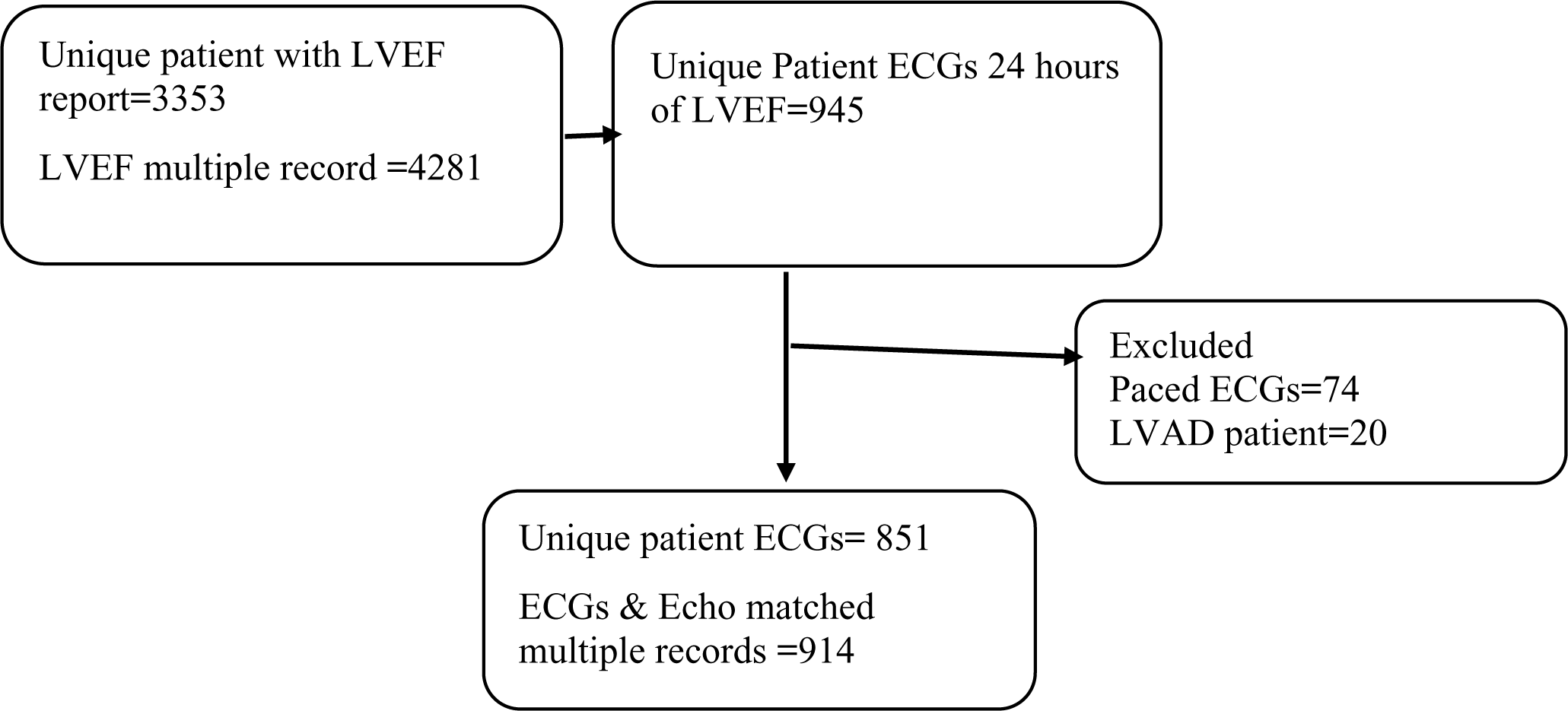
Sample Selection for this Study.

### Data Extraction

We collected data through the University of Rochester’s Clinical and Translational Science Institute (CTSI); a team of nurses and data management specialists extracted the electronic health record data using ICD-10 codes of patients with an ECG and echo report within 24 hours span to maintain the clinical stability of data. We requested clinical data such as demographics (age, race, sex), smoking status, vital signs closest to time of the echocardiogram (body mass index, blood pressure, respirations), comorbidities based on ICD code-10, inpatient HF medications (including angiotensin-converting enzyme inhibitors, beta-blockers, mineralocorticoid receptor antagonists, angiotensin receptor blockers, and diuretics), ECG and echocardiogram report.

### ECG Data

At our institution, we acquired ECG data correlating with the availability of LVEF records. We recorded these ECGs using the Philips TC70 cardiograph while patients were supine. We downloaded the automated interpretation of these ECGs, excluding those with noise and artifacts filtered with 159Hz bandwidth for maximum fidelity and sampling rate at 500 samples per second per electrode/lead. The automated interpretation exhibited 87% of sensitivity and 99% of specificity. ^17^ Each 12-lead ECG captured data for 10 seconds, digitalized at 500 samples per second, with a recording speed of 25 millimeters per second (mm/s) and a standard calibration of 10 mm/millivolts(mV). We electronically stored these ECGs in the ItelliVue ECG management database (Philips Healthcare) and linked them to patient records via medical record numbers and hospitalization dates^18,19^. We obtained PDF copies of the morphology and analysis performed by the Philips DXL advanced algorithm and extracted measurements. We converted them into a digital, analyzable comma-separated values (CSV) format using Python code integrated with optical character recognition (OCR) software from Nanonets. This custom code is available on GitHub for public access^20^.

We found 3353 patients with echocardiogram reports; only 30% (n=994) patients had ECG records in Philips within 24 hours of the echocardiogram report. To ensure data accuracy upon downloading the ECGs in PDF format, two investigators systematically reviewed them for correct lead placement and missing data due to artifacts, identifying 80 ECGs with missing data due to artifacts. Total ECG morphology involved 527 features with less than 10% missing values, had these values replaced with the mean of other leads reflecting the same aspect of the heart to ensure completeness, consistency, and accuracy (Figure 1).

### Outcome Variable

We aimed to estimate LVEF ≤30% in AHF patients; therefore, we categorized LVEF % into ≤30% and >30%, for model performance. We collected all the LVEF reports only from the echocardiogram to ensure consistency among the measurements ^21^.

### Statistical Analysis

We presented the patients’ characteristics as counts, percentages, means, medians, standard deviations, and interquartile ranges as appropriate. We trimmed less than 10% outliers with a 95% confidence limit boundary. To select the important variables in the ≤30%LVEF prediction model, we used the exploratory method Least Absolute Shrinkage and Selection Operator (LASSO) using the “cv. glmnet” function from the “glmnet) Packages in R Programming language, which implemented logistic regression with LASSO regularization^22^.This method applied a penalty to the absolute size of the regression coefficients, effectively shrinking some of them to zero, simplifying the model, and selecting only the most relevant predictors. The degree of regularization was determined by the parameter “lambda,” which controlled the trade-off between fitting the data and keeping the model simple. We split data in a 70:30 ratio into training and testing groups, allocating 70% for training our predictive model and 30% for testing its performance. To optimize the selection of the “lambda” parameter, we used 5-fold cross-validation ^23^. This approach divided the dataset into five subsets, using four subsets for training the model and the remaining one for the validation, iteratively. The “cv.glmnet” function automatically performed this cross-validation, selecting the “lambda” value that minimizes the cross-validated error. The final model was determined at the “lambda” value that resulted in the minimum mean cross-validated error (“lambda. min”)^24^. Coefficients of the predictors in the final model were extracted, allowing us to identify the most predictive variables contributing to the ≤30%LVEF prediction. The coefficients of the important variables were displayed, indicating their respective contributors to the model. Predictors with non-zero coefficients were considered important, revealing their relevance in the context of estimating LVEF.

### LASSO Regression

We used LASSO to identify ECG features in LVEF estimation. LASSO regression is particularly well-suited for selecting key features in a model that estimates LVEF from a large set of ECG data due to its inherent property of feature selection ^23^. LVEF estimates involve analyzing a potentially large set of variables from ECG readings, many of which may be irreverent or redundant. LASSO, with its L1 regularization technique, not only helps in avoiding overfitting by penalizing the absolute size of the coefficients but also effectively shrinks the less important feature coefficient to zero, thus excluding them from the model. ^25^ This automatic feature selection simplified the model, making it easier to interpret and faster to compute, which is crucial in clinical settings for timely decision-making. By focusing only on the most significant predictors, LASSO regression enhances the model’s predictive accuracy while maintaining simplicity, making it an excellent choice for constructing a robust and interpretable model for estimating ejection fraction from ECG data ^22^.

Using the optimized Lasso regression, we predicted the outcomes from our dataset. The “predict” function in R was used to generate predicted probabilities (“lasso_pred”) for each observation in the dataset, utilizing the model parameters determined at the optimal “lambda.min” value. Furthermore, we constructed a confusion matrix to evaluate the model’s performance, tabulating actual versus predicted classifications. This matrix provided a straightforward visualization of the model’s performance, including the counts of true positives, true negatives, false positives, and false negatives. The confusion matrix was further converted into a heatmap using the “ggplot2” package in R. This graphical representation allowed us to visually assess the model’s performance, with different color intensities and the frequency of each outcome category (e.g., true positives, true, negatives).

In addition, to quantitatively assess the model’s discriminative ability, we plotted receiver operating characteristics (ROC) curves and AUC. The ROC curve, created using “pROC” packages in R, plotted the true positive rate against the false positive rate at two different threshold settings. A valid positive test (true positive) is most important for cost-effectiveness ^26^. Therefore, the cutoff threshold for binary prediction of ≤ 30% was set at 0.10 based on threshold that achieved a sensitivity of >90% and expert’s preferences^27^. Another approach was using Youden method to balance between positive and negative cases which selects the optimal threshold value or cutoff point ^28^. Further we evaluated area under curve (AUC), sensitivity, specificity, accuracy, and negative predictive value (NPV).

The LASSO approach identifies important variables but does not provide causal interpretations. To understand how each predictor is associated with the probability of the outcome, the logistic regression model was fitted using the “glm” function in R. We obtained a detailed summary of the fitted model, including the coefficients for each predictor, and *p*-values. We computed 95% confidence intervals for each coefficient using the profile likelihood method. The analysis was conducted in RStudio, using a statistical significance cut-off of a two-tailed *p*-value of <0.05.

## Results

### Demographic and Clinical Characteristics

The demographic and clinical characteristics of the study population are presented in Table 1. The study population comprised 815 patients, with ≤30% LVEF a subgroup of 16.45% (n=140) patients identified as having ≤30% LVEF. The median age of the total cohort was 74 years, with interquartile range (IQR) of 11, while the median age within the ≤30% LVEF group was at 74, with a wider IQR of 19. Males constituted a larger proportion of both the total patient group (56%, n=478) and the ≤30% LVEF subgroup (61%, n=86). In terms of racial distribution, 91% (n=771) of the total cohort were White. A significant majority of the total cohort (34%, n=198) and the subgroup (73%, n=102) were current or former smokers.

**Table 1.**
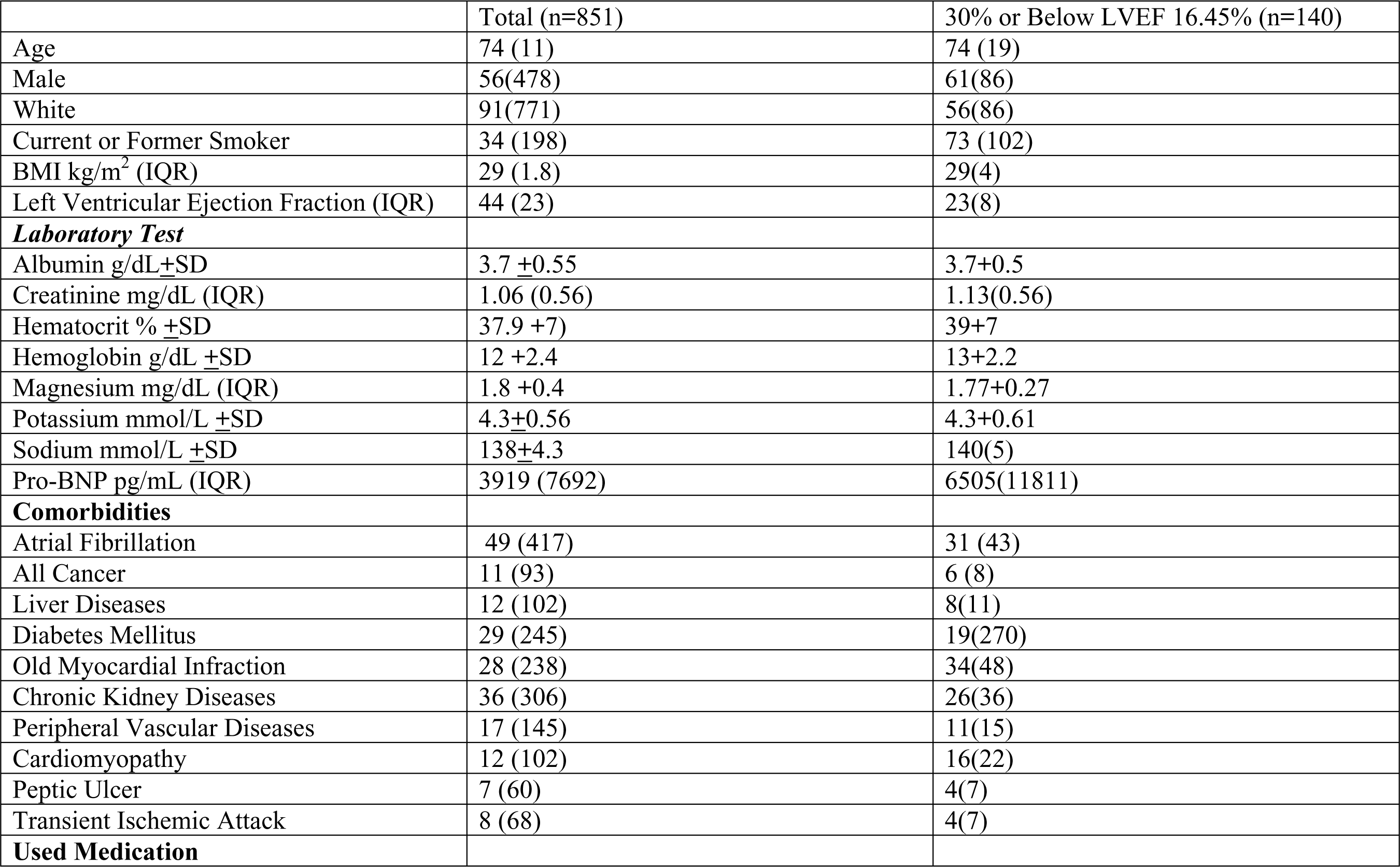
Demographic and Clinical Information N=851.

The median body mass index (BMI) was 29 kg/m2(IQR of 1.8) for the total cohort and similar for the subgroup. The prevalence of comorbidities was reported, with history of atrial fibrillation 49% (n=417), previous myocardial infarction 28% (n=238) and chronic kidney diseases (36%, n=306) being the most common in the total cohort. Among patients with ≤30% LVEF, atrial fibrillation (31%, n=43), old myocardial infarction (34%, n=48), and chronic kidney diseases (26%, n=36) were also prevalent. Laboratory tests revealed that the median Pro-BNP in all patients were 3919pg/mL (IQR of 7,692), showing notably higher level (6,505 pg/mL) in ≤30% LVEF group.

We collected multiple ECG and echocardiography records from 851 patients which counts a total of 914. Among 914 ECGs 21% (n=191) had atrial fibrillation/flutter, 3% (n=49) presented left ventricular hypertrophy (LVH), 7% (n=62) with left bundle branch block (LBBB), and 5% (n=50) showed right bundle branch block (RBBB). Median ECG to EF measurement time was 9 hours (IQR of 9 hours) (Table 2).

**Table 2.**
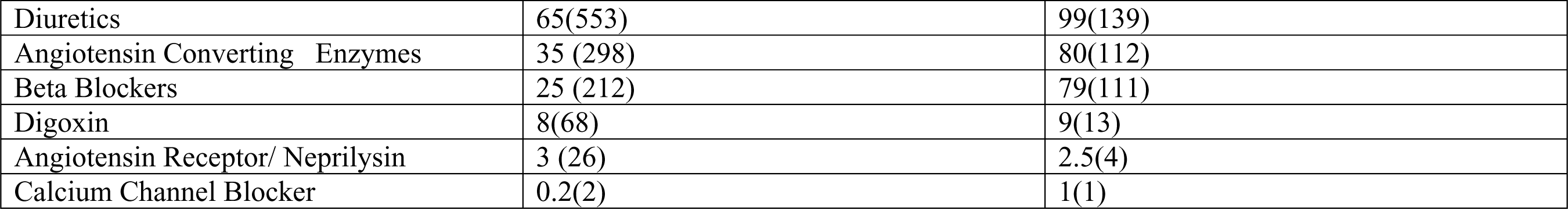
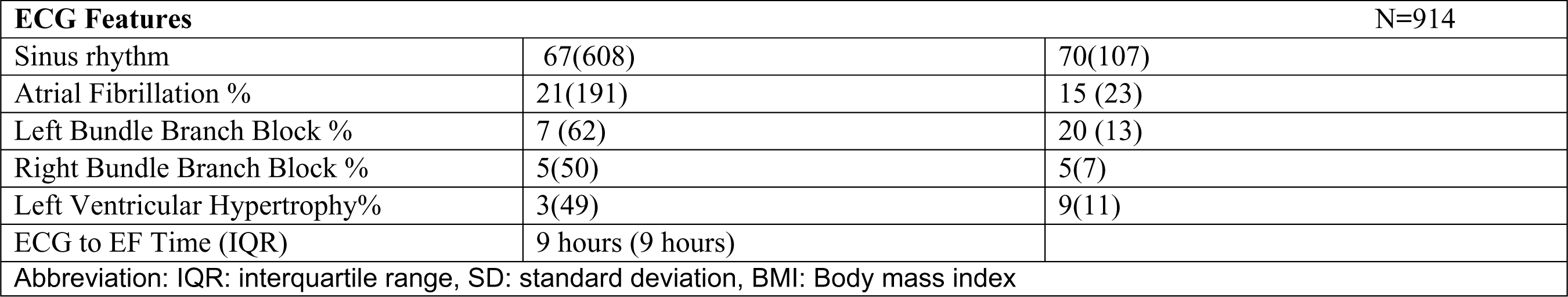
ECG Features.

### Model Performance for ≤30% LVEF

To evaluate the model performance, we used multiple records of ECGs within the ECG to echocardiogram 24 hours window period and all together 914 ECGs and echocardiograms were matched. Among the total 527 ECG features, the Lasso model demonstrated 44 (8%) ECG features as important features for estimating ≤30% LVEF. We examined and removed features with very high collinearity scores that contain redundant information (for example, we kept mean QTc interval if the model selected both QT and QTc. We removed two ECG features were QT interval in II and aVR, 1finally 42 remained the important ECG features (Figure 2). We evaluated model performance with or without P waves, because 21% (n=191/914) ECGs had atrial fibrillation/flutter which do not present the measurement of P waves. But, the model performance with P waves performed better compared to without P waves. The lollipop chart in Figure 2 visualizes the coefficient of the predictions derived from LASSO regression model. Precordial or chest leads on lateral wall of the left ventricle showed the highest positive and negative coefficient. The ST segment mid at V4 & V5 and ST end at V5, the terms relate to measurements at the midpoint and end of the ST segment in lead V4 and V5 that gives the horizontal view of the heart’s electrical activity, particularly the anterior and lateral regions of the left ventricle had the most significant negative coefficient, suggesting a strong negative impact on the outcome.

**Figure 2.**
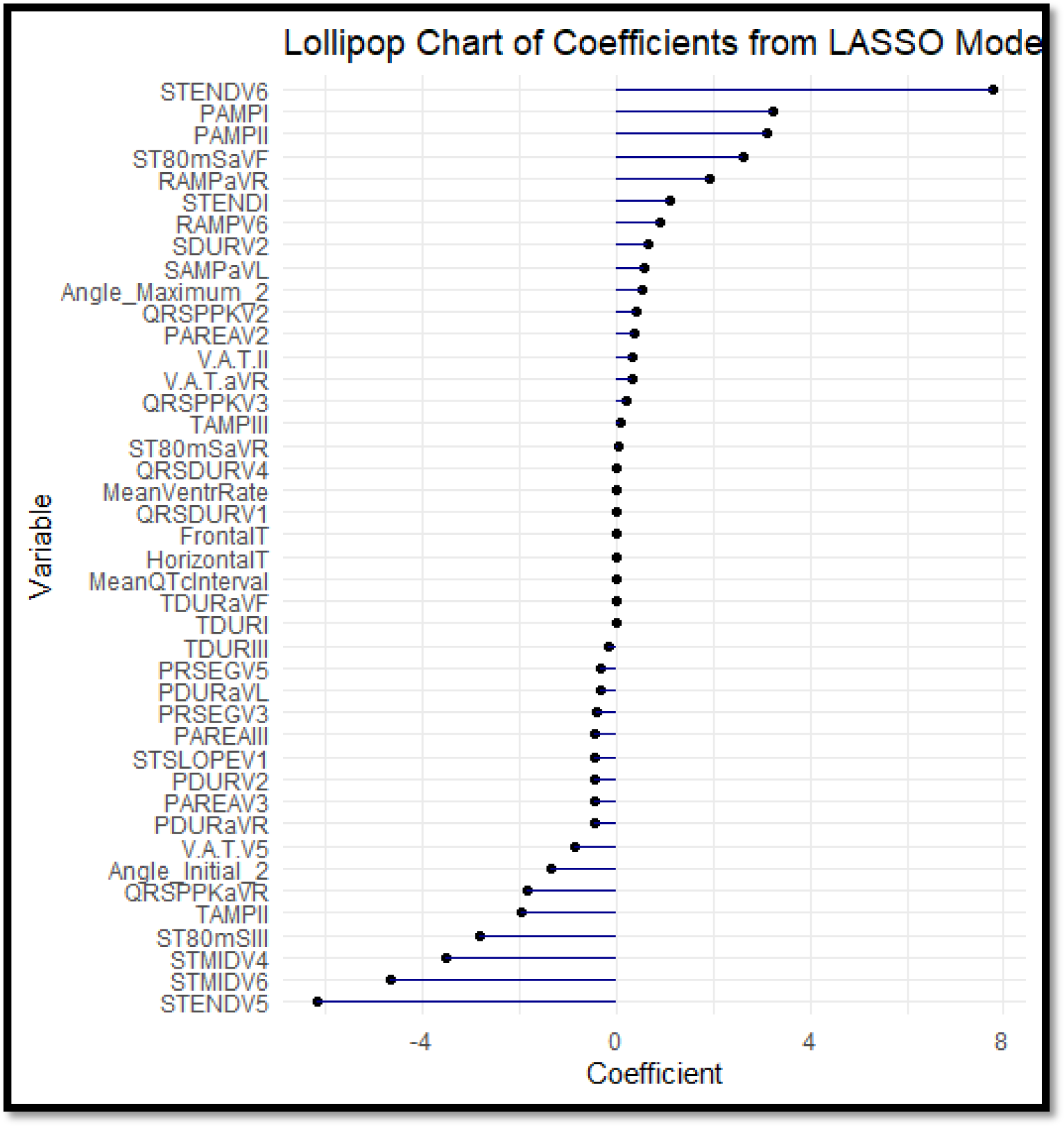
Important Variable Selection by LASSO for ≤30% LVEF Estimation: The figure presents the coefficients of the predictors’ variables identified as significant in the LASSO regression model. Each bar represents the magnitude and direction of the association between the predictor variables (displayed on the horizontal axis) and the response variable, with the length of the bar indicating the strength of the association. Positive coefficients are shown extending right and suggest a positive relationship with the outcome, while negative coefficients extend left and indicate an inverse relationship. Variables with longer bars have a greater impact on the model, either increasing (if positive) or decreasing (if negative) the likelihood of the ≤30% LVEF Estimation. The intercept, or the baseline level of the outcome when all predictors are zero, is also shown.

Logistic regression for estimating ≤30% LVEF showed varying association by several ECG leads. Increase in initial axis of the QRS complex indicating the early ventricular depolarization (Estimate: −1.464, 95% CI: −2.97 to −0.391, *p*=0.027), peak amplitude of R wave in the QRS complex in lead aVR (Estimate: −1.89, 95% CI: −2.92 to −0.90, *p*=<0.0001), more negative deflection in ST segment at lead V5 (Estimate: −6.67, 95% CI: −10.06 to −3.46, *p*=<0.0001) and increase in ST slope in aVR (Estimate: −0.46, 95% CI: −0.78 to −0.15, *p*=0.004) decreases the odds of ≤30% LVEF. Likewise, delayed in ventricular activation time in lead V5 are likely to be in ≤30% LVEF (Estimate: −0.91, 95% CI: −1.48 to −0.35, *p*=0.002).

Conversely, increase in maximum angle increases likelihood of <u><3</u>0% LVEF (Estimate: 0.56, 95% CI: 0.29 to 0.84, *p*=<0.0001). Increase in mean ventricular rate (Estimate: 0.12, 95% CI: 0.001 to 0.024, *p*=0.038)., increased peak amplitude of the R wave in the QRS complex in lead V2 (Estimate: 0.44, 95% CI: 0.09 to 0.78, *p*=0.01), increase R amplitude in lead aVR and V6 (Estimate: 2.08, 95% CI: 0.09 to 4.03, *p*=0.03) and (Estimate: 0.94, 95% CI: 0.34 to 1.56, *p*=0.002), higher ST end at V6 (Estimate: 9.85, 95% CI: 4.09 to 16.03, *p*=0.001) increases the higher odds of ≤30% LVEF (Table 3).

**Table 3.** Logistic Regression for Estimating for ≤30% LVEF.

| Term | Estimate | Lower 95% CI | Upper 95% CI | Pr(>\|z\|) |
| --- | --- | --- | --- | --- |
| (Intercept) | 1.313 | -5.253 | 7.931 | 0.696 |
| Angle Initial | -1.464 | -2.970 | -0.391 | 0.027 |
| Angle Maximum | 0.562 | 0.291 | 0.849 | 0.000 |
| Frontal T | 0.003 | -0.000 | 0.007 | 0.075 |
| Horizontal T | 0.002 | -0.002 | 0.007 | 0.341 |
| Mean QTc Interval | 0.002 | -0.004 | 0.009 | 0.500 |
| Mean Ventricular Rate | 0.012 | 0.001 | 0.024 | 0.038 |
| P amplitude I | 3.488 | -1.022 | 8.052 | 0.131 |
| P amplitude II | 3.125 | -0.490 | 6.914 | 0.098 |
| P amplitude III | -0.443 | -0.917 | 0.035 | 0.067 |
| P AREA V2 | 0.383 | -0.071 | 0.879 | 0.112 |
| P AREA V3 | -0.474 | -0.974 | 0.033 | 0.064 |
| P duration aVL | -0.359 | -0.851 | 0.132 | 0.152 |
| P duration aVR | -0.487 | -1.088 | 0.123 | 0.114 |
| P duration V2 | -0.469 | -0.990 | 0.043 | 0.075 |
| PR segment V3 | -0.425 | -0.853 | 0.006 | 0.051 |
| PR segment V5 | -0.341 | -0.714 | 0.043 | 0.076 |
| QRS duration V1 | 0.012 | -0.002 | 0.028 | 0.101 |
| QRS duration V4 | 0.015 | -0.001 | 0.031 | 0.065 |
| QRS PPK aVR | -1.894 | -2.922 | -0.908 | 0.000 |
| QRS PPK V2 | 0.441 | 0.095 | 0.788 | 0.012 |
| QRS PPK V3 | 0.194 | -0.186 | 0.569 | 0.313 |
| R amplitude aVR | 2.083 | 0.096 | 4.033 | 0.037 |
| R amplitude V6 | 0.947 | 0.341 | 1.561 | 0.002 |
| S amplitude aVL | 0.616 | 0.004 | 1.498 | 0.101 |
| S duration V2 | 0.693 | -0.074 | 1.505 | 0.085 |
| ST80mS aVF | 2.926 | 0.519 | 6.128 | 0.060 |
| ST80mS aVR | 0.186 | -2.231 | 1.253 | 0.770 |
| ST80mS III | -2.886 | -7.322 | 1.366 | 0.192 |
| ST END I | 1.184 | 0.242 | 4.276 | 0.114 |
| ST END V5 | -6.669 | -10.069 | -3.468 | 0.000 |
| ST END V6 | 9.854 | 4.095 | 16.037 | 0.001 |
| ST MID V4 | -3.939 | -10.347 | 2.298 | 0.221 |
| ST MID V6 | -7.041 | -14.343 | -0.857 | 0.052 |
| ST SLOPE V1 | -0.466 | -0.781 | -0.153 | 0.004 |
| T amplitude II | -2.240 | -4.619 | -0.317 | 0.062 |
| T amplitude III | 0.078 | -0.677 | 0.854 | 0.842 |
| T duration aVF | -0.002 | -0.007 | 0.002 | 0.267 |
| T duration I | -0.003 | -0.007 | 0.001 | 0.090 |
| T duration III | -0.174 | -0.509 | 0.185 | 0.316 |
| Ventricular activation time aVR | 0.326 | -0.093 | 0.778 | 0.139 |
| Ventricular activation time II | 0.320 | -0.265 | 0.922 | 0.290 |
| Ventricular activation time V5 | -0.914 | -1.488 | -0.356 | 0.002 |

The ECG to EF ≤30% provided a high degree discrimination AUC of 0.86, 95% confidence interval (CI) 0.83 to 0.89. Using a threshold to yield a 90% sensitivity on the test data, the sensitivity was 92 (95% CI: 88% to 96%), specificity 53% (50% to 57%), overall accuracy 60% (95% CI:60 % to 63%) and, negative predictive value of 95%. Furthermore, we tested the model performance in training data. The AUC in training data shows 0.87 (95% CI: 0.83 to 0.90), sensitivity increased to 97% (95% CI: 91% to 99%), specificity of 53 % (95% CI: 48% to 57 %), and accuracy of 61% (95% CI: 61% to 65%) and negative predictive value of 99%. When selecting a threshold with no preferences for sensitivity, the overall accuracy was 0.86, with a sensitivity of 85% (95% CI:0.68 to 90), the specificity was 67% (95% CI:0.66 to 87%), with 74% accuracy (95% CI: 71% to 77%) (Figure 3). Youden method in training shows AUC of 0.87 (95% CI: 0.84 to 0.91), sensitivity 84% (95% CI: 67% to 94%), specificity of 74% (95% CI:63% to 91%) with optimal threshold of 0.16 (Figure 4) (Figure 5).

**Figure 3.**
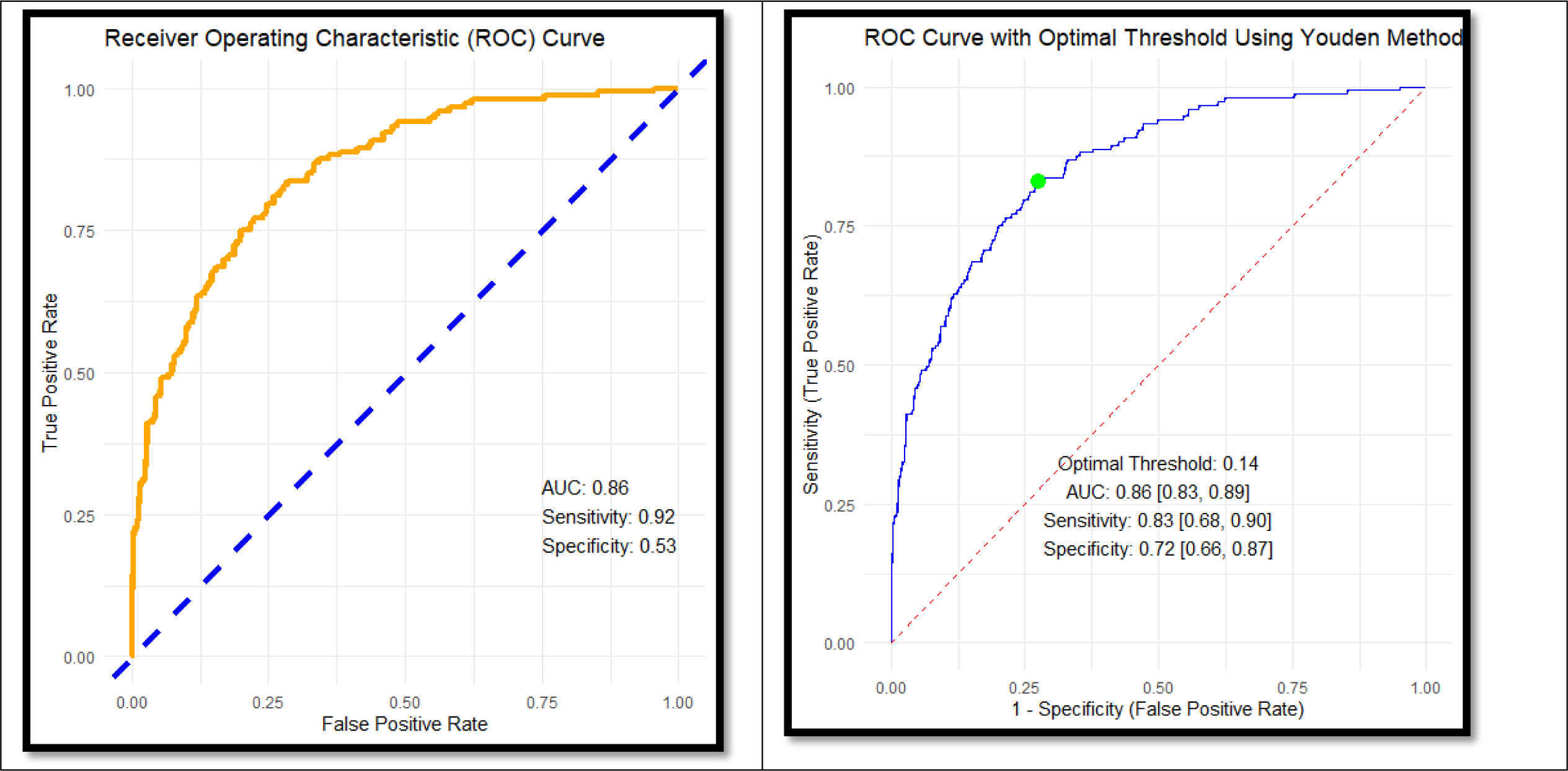
ROC for ≤30% LVEF estimation model: The ROC curve plot on left side shows the sensitivity (true positive) on the y-axis versus 1-specificity (false positive rate) on the x-axis yielding 90% sensitivity, and right side shows the Youden method model performance.

**Figure 4.**
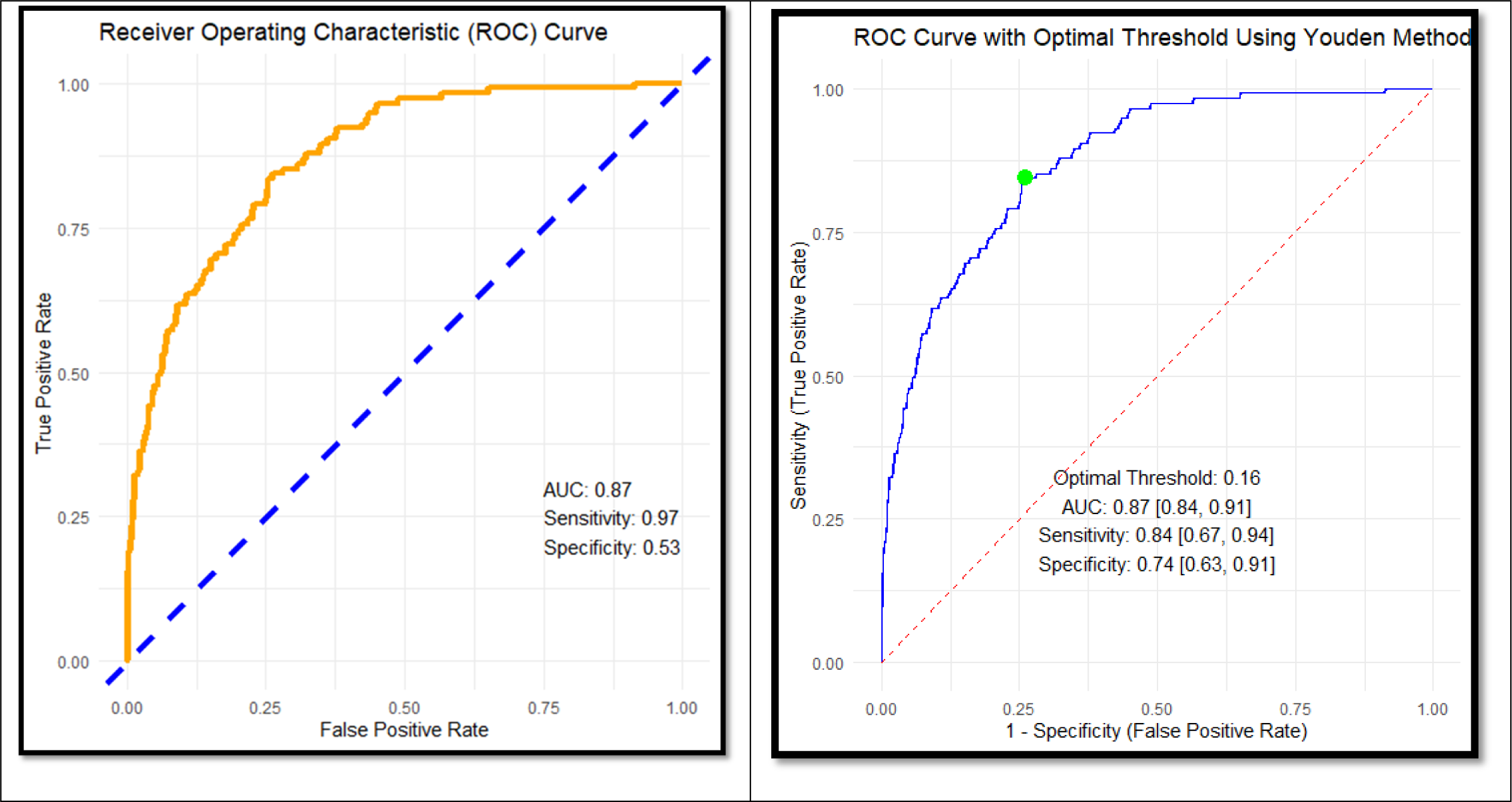
ROC for ≤30% LVEF estimation model using training data: The ROC curve plot on left side shows the sensitivity (true positive) on the y-axis versus 1-specificity (false positive rate) on the x-axis yielding 90% sensitivity, and right side shows the Youden method model performance.

**Figure 5.**
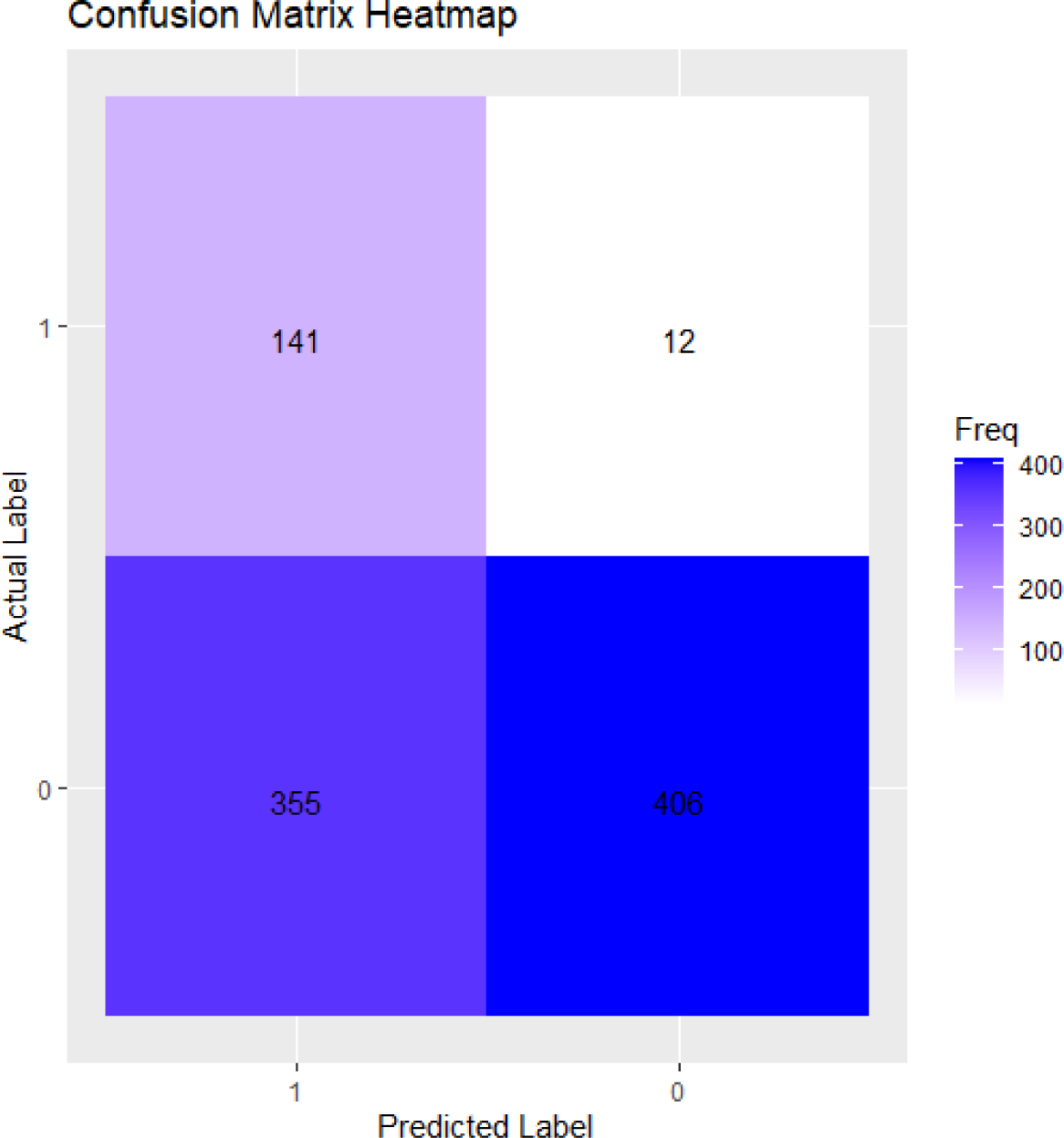
Actual versus Predicted ≤30% LVEF estimation: Confusion Matrix Heatmap of Predicted vs. Actual Labels: The heatmap illustrates the performance of the classification model. Each cell represents the count of observations for the actual label (vertical axis) versus the Predicted label (horizontal axis). The color intensity represents the frequency of observations, with darker shades indicating higher frequencies. The top left quadrant of the matrix shows the number of true positives (TP), which in this case is 141. The bottom right quadrant shows the number of true negatives (TN), here being 406, indicating the cases where the model correctly predicted the negative class. Conversely, the top right quadrant contains the false negative (FN), numbering 12, where the model incorrectly predicted the negative class when the actual label was positive. The bottom left quadrant represents the false positive (FP), which is 355, showing the cases where the model incorrectly predicted the positive class.

## Discussion

This is a comprehensive evaluation of automated 12-lead ECG features to estimate LVEF in patients who underwent both ECG and echocardiogram assessment during hospitalization within 24 hours. Utilizing the Lasso model, we identified 42 important features contributing to estimating ≤30% LVEF. Notably, several features positively associated with ≤30% LVEF, including maximum angle, mean ventricular rate, peak to peak QRS in lead V2, R amplitude in aVR and V6, and ST end in V6. And initial angle, peak to peak QRS in aVR ST end in V5 and V6, ST slope in V1 and ventricular activation time in V5 were inversely associated to estimate ≤30% LVEF. These findings illuminate the critical role of specific ECG markers in signifying reduced LVEF.

The diagnostic model was evaluated using dataset of 914 matched ECGs and echocardiographic records. Overall, the model demonstrated a high sensitivity of 91%, indicating a strong ability to correctly identify true positive cases. However, specificity was low at 54%, suggesting a moderate rate of false positives. Numerous investigations have utilized artificial intelligence for predicting EF, with some targeting asymptomatic left ventricular dysfunction ^8,14^ and HF^12,13,29^. In our study, we dichotomized ≤30% LVEF and above, yielding an AUC of 0.86, a sensitivity of 91%, and a specificity of 54%. Conversely, a comprehensive study conducted at Mayo Clinic, encompassing a vast sample size (625,326), utilized a neural network within a comparable ECG to echocardiogram window period, reporting lower sensitivity (86% vs. 91%) and higher AUC (0.93 vs 0.86) than our findings.^8^ Our utilization of LASSO regression, inherently selecting a parsimonious set of influence ECG variables, might be a key reason for the observed performance disparities. This method tends to set model parameters for LVEF that are unlikely to significantly predict the observations to zero, thereby effectively excluding them from the regression model. This selection deliberates the selection process and acts as a safeguard against overfitting. Additionally, notable distinctions exist between our study and theirs. Their sample comprised a younger population, focused on asymptomatic left ventricular dysfunction compared to our cohort. Notably, the majority (89%) of LVEF measurements were obtained within one day of the index ECG^8^ in their study, but we included 24 hours ECG to EF time.

In a study in a Korean population focusing on HF (with a sample of 22,765), a slightly lower AUC (0.84) was observed than ours, utilizing a longer ECG to echocardiogram window of 4 weeks through artificial intelligence^12,13^. The long duration between window ECG and echocardiogram assessment likely contributes to errors associated with temporal differences in these findings. Moreover, Chen et al. (2020) investigated ECG to echocardiogram interval within a week time, on average three days, longer than our observed average ECG to echocardiogram time (typically <1 day). Their study utilized AI models based on baseline ECG features studied within asymptomatic left ventricular dysfunction^14^. It is important to note that AI models, particularly deep and neural models, are often considered “black boxes.” Despite their effectiveness in prediction, these models lack interpretability in deciphering the decision-making process leading to clinical endpoints^13^.

The utilization of ECG features for LVEF prediction exhibits variation across studies. For instance, Alhmayedeh et al. (2020) analyzed 554 automated 12-lead ECG features to predict LVEF in suspected acute coronary syndrome. They employed multivariate linear regression analysis and the regression tree with the deviance method to rank the most important ECG features for estimating reduced LVEF. However, their study reported lower model performance (45.2%) than our findings. It is important to note that this study had a relatively small sample size, which might have limited the statistical power of their findings. Additionally, the authors do not report the time interval between ECG and echocardiogram, an important factor that could influence model performance^30^.

LASSO regression is a pivotal tool in developing accurate LVEF prediction models. Its utility lies in the ability to discern significant variables for each LVEF group, effectively averting model overfitting. By identifying distinct important variables for each LVEF group, this approach contributes to more robust and reliable model development. Our study’s findings indicate promising prospects in predicting reduced LVEF among patients. The highlighted high sensitivity in predicting reduced LVEF is paramount in the clinical triage process, particularly within EDs. High sensitivity in a diagnostics model is of substantial clinical and economic benefits^26^. From the economic standpoint, this characteristic is advantages as it reduces the need for repeat testing. Missing a diagnosis often leads to additional tests, which not only increases healthcare costs but also delays critical treatment, potentially exacerbating the condition and leading to more expensive interventions later on. Furthermore, early detection testing can decrease the likelihood of complications that require more complex and costly care. By ensuring that most patients with the conditions are identified quickly, the model aids in the timely initiation of treatment. This early intervention can prevent the progression of the condition, which in turn can lead to substantial cost savings by reducing the need for more extensive medical procedures and long-term care^27^.

This study’s detailed ECG features analysis lays the groundwork for creating advanced tools for heart function assessment. Key ECG indicators, linked to ≤30% LVEF, could improve algorithms for detecting patients at risk. These indicators are especially useful for preliminary screenings in settings lacking echocardiography aiding early cardiac care decisions. Additionally, our finding could enrich AI cariology models with precise data for LVEF prediction, offering clinicians real-time diagnostic support.

Three primary limitations warrant consideration in our study. Firstly, the retrospective nature of the study, conducted within a single institution, raises the need for further validation through prospective studies to ascertain the accuracy and clinical applicability of ECG to EF predictions. In the future, it is also important to study how such results affect HF outcomes after clinicians see these results. Although ECG data were collected from multiple hospital sources within the institution, validating our findings across diverse cohorts and healthcare settings remains an area for future investigation. In addition, only 38% of the patients had ECG records available in the ECG management database. This limitation has several implications for both the generalizability and the validity of our findings which could bring from selection bias.

Secondly, an important point to acknowledge is the non-concurrent ECG and echocardiogram data collection. While we limited time within 24 hours, ECG changes may occur in response to treatment interventions. The temporal variance between our study’s ECG and echocardiogram recordings may introduce a potential temporal bias in our findings, influencing the observed relationship between ECG features and EF estimation.

Furthermore, it is noteworthy that our patient’s cohort primarily consisted of individuals hospitalized for symptom management. Our findings might have limited generalizability to populations with non-symptomatic or stable. Considering the varying clinical presentations across AHF severity levels, it is essential to exercise caution when extending our results to these distinct subsets of patients.

## Conclusions

In conclusion, our study presents a comprehensive analysis of ECG features to predict reduced LVEF, highlighting the potential of automated 12-lead ECG analysis in enhancing diagnostic accuracy for heart failure. By employing the LASSO model, we identified key ECG features that significantly contribute to the estimation of ≤30% LVEF, with the model demonstrating a high sensitivity and moderate specificity. The study also acknowledges the limitations posed by the retrospective design and the need of validation in broader, prospective, and clinical trial populations. Nonetheless, our results offer promising insights into the utility of ECG-based diagnostic tools and artificial intelligence in cardiology, potentially paving the way for more accurate, real-time assessment of cardiac function and risk stratification in clinical settings, which could positively impact patient outcomes.

## Data Availability

Data sharing is reserved to a corresponding author

